# Patient Perspectives on Potential Implementation of Coordinated Family Care Visits for Inherited Cardiovascular Disease

**DOI:** 10.64898/2026.08.05.26359830

**Authors:** Emma Draisin, Hafsah Badar, Hetanshi Naik, Julia Platt, Beth Kaufman, Heidi Salisbury, Hannah Ison

## Abstract

**Introduction:** Shared medical appointments (SMAs) are medical visits where multiple individuals are seen together in a group setting. For patients with inherited cardiovascular disease, where multiple family members often require ongoing cardiac care and screening, family SMAs may be particularly valuable as a tool to facilitate family communication and comprehension of their condition. This research aimed to identify patient perspectives on the potential benefits and challenges of family SMAs in comparison to an existing individual clinic model.

**Methods:** Qualitative semi-structured interviews were conducted with adult family representatives. Each family had at least one family member seen at the adult and pediatric inherited cardiovascular disease clinics. Interview recordings were transcribed verbatim and inductively coded using a content analysis approach.

**Results:** Sixteen families were interviewed in this study. The mean age of the family representative interviewed was 43.4 years (± 9.3 SD), and they were followed at Stanford Health Care for a mean of 7.3 years (± 4.2 SD). 81.2% (13/16) of families said they would find family SMAs beneficial. For interested families who consented to recorded interviews (n=12), benefits and challenges fell into two major categories: care quality and access and logistics. Interested families thought family SMAs would provide an added care quality benefit by increasing understanding among adults, children, and providers (83.3%, 10/12). Six of twelve participants interested in having family SMA visits felt there would be logistical/access-based benefits to this new model (50%, 6/12). Families also identified possible challenges with this model, such as less individualized care, potential privacy concerns, and concerns regarding the smoothness of the clinic process in coordinating a family SMA.

**Conclusion:** The majority of families believed a family SMA model would provide added benefit to families with inherited cardiovascular disease, but requires thoughtful implementation and should be tailored to families’ unique needs.

## Introduction

Inherited cardiovascular diseases require longitudinal, multigenerational care that spans pediatric and adult cardiology, with integrated genetic counseling and psychosocial support.^1,2^ Evidence suggests that patients with genetic conditions may benefit from family-centered care (FCC).^3^ FCC is a model of care that describes planning around the whole family by integrating family-centered values of information sharing, shared decision-making, family involvement, and family education, support, and training.^4,5^ Additionally, FCC incorporates collaboration between parents and the multi-disciplinary medical team, and emphasizes health care that is flexible, culturally sensitive, and adaptable to individual family needs.^6^

FCC has been adopted as a care model in many pediatric healthcare systems.^7,8^ To date, the focus of FCC literature has been primarily on pediatric patients and complex patients who require the assistance of caregivers. Systematic reviews of FCC outcomes have shown improvements for patients quality of life, knowledge/comprehension, and satisfaction with care.^5,9–11^ Affected family members have also reported improved outcomes of knowledge, confidence, and family functioning.^5^ While FCC has been shown to have many positive outcomes, family needs for coordination of care are not met by the FCC model alone. Parents of pediatric patients have shared frustration that despite the FCC model, there is still insufficient communication between providers, leading to challenges in scheduling appointment times and coordinating multidisciplinary care.^11^

One care model that could benefit families with inherited conditions is shared medical appointments (SMAs). SMAs are group visits of multiple individuals impacted by the same condition. This model of care has gained popularity in both primary care and specialty settings with the goals of improving access, enhancing patient education and self-efficacy, and reducing costs.^12,13^ Systematic reviews on experiences of SMAs show that it can improve patient perception of quality of care and quality of life.^12,13^ There is variability in how SMAs are structured in different healthcare systems, and reports on the impact of SMAs on time and cost efficiency are mixed.^12,13^

Studies on the use of SMAs in the family setting, with multiple family members having the same condition, is very limited. There have been a few reports of clinics offering group appointments to families navigating inherited genetic conditions, primarily in order to organize cascade genetic testing once one family member has been identified to have a pathogenic variant.^14–17^ One recent study examined genetic counselor opinions on family group appointments, and found that benefits included promoting familial support and communication.^14^

To our knowledge, globally, there are two clinics that have publicized providing family SMAs for cardiovascular genetic disorders, with one limited to inherited arrhythmias.^17,18^ Given that inherited cardiovascular conditions are often inherited in an autosomal dominant manner, placing first-degree family members of affected individuals at increased risk, and often requiring both children and parents to receive ongoing cardiac follow-up and screening, family SMAs that incorporate FCC principles may help improve information sharing, family communication, and family support. This study aimed to better understand patient perspectives on the potential benefits and challenges of family SMAs for inherited cardiovascular disease.

## Methods

### Study Design

To identify patient perspectives, we conducted interviews with families impacted by inherited cardiovascular disease to understand their experience with the existing clinic model whereby parents and children are seen in separate visits, and their perspectives on family SMAs. We conceptualized family SMAs as appointments with multiple family members and a team of a pediatric cardiologist, an adult cardiologist, an advanced practice registered nurse, and a genetic counselor. We used a mixed methods study design consisting of inductive content analysis on qualitative semi-structured interviews, and quantitative analysis of Likert scale questions.

### Participant sample

Interviews were conducted with adult family members acting as family representatives. Each family had at least one family member seen at the Stanford Center for Inherited Cardiovascular Disease (SCICD) and at least one family member cared for in the Pediatric Inherited Cardiovascular Disorders program (PICD). Eligible families were identified by study staff who were part of the families’ care teams. All family representatives were 18 years of age or older, English-speaking, and had been previously seen by the team. Initial interviews (n=14) were conducted by a study team member (HB) between May 2024 and June 2024, and additional interviews (n=2) were conducted by a second study team member (ED) between October 2025 and November 2025. Informed consent was obtained from all participants. This study was approved by the Stanford University Institutional Review Board (IRB # 45129).

### Data collection

Interview questions were developed to understand family experiences with current care at both SCICD and PICD, and to assess interest in family SMAs. A group of experts were convened to discuss and develop the questions via a committee consisting of senior staff and experts across the multidisciplinary team, as well as patient advisors. The interview guide consisted of four 5-point Likert-scale questions and six open-ended questions (see supplementary material). Demographic information collected for each family included the age and sex of adult and pediatric family members, as well as the relationship of each family member to the family representative interviewee. Family representative demographic information abstracted from the EHR included age, years of follow-up at SCICD, self-reported race/ethnicity, assigned sex at birth, clinical indication, and home distance to clinic. Interviews were conducted via Zoom by two study staff members who were not involved in the clinical care of any patients interviewed. Family representatives were asked to consent to the recording of their interview; n=14 consented and n=2 abstained and opted for interview notes only. Recorded interviews and interview notes were stored in a PHI-safe drive. Interview recordings were transcribed verbatim by a HIPAA-compliant transcription service, and manually checked by a research team member (ED). Once adequate information power^19^ was deemed achieved, data collection was considered complete.

### Data analysis

Recorded interviews (n=14) were inductively coded in NVivo software using a content analysis approach^20^ to categorize benefits and challenges identified in both the current clinic model and the proposed family SMA model. Inductive content analysis was selected as it is well suited to practice-oriented research and can support actionable recommendations for implementation.^20^ One coder (ED) reviewed all initial transcripts and developed a preliminary codebook, which was reviewed and modified by members of the research team. Code refinement continued throughout the analysis process. Category development focused on identifying benefits and challenges shared by participants. For Likert-scale questions, data were analyzed quantitatively and summarized using descriptive statistics.

## Results

Sixteen families were included in this study. The mean number of adult patients with inherited cardiovascular disease per family was 1.9 (range 1-3), and the mean number of pediatric patients per family was 2.1 (range 1-5) (Table 1). Within each family, one adult member was chosen as the family representative and interviewed. Six of sixteen interviewed family representatives were the index patient, which was defined for the purpose of this study as the first person in the family seen at Stanford for inherited cardiovascular disease. Of the remaining family representatives, 2/16 were the child, 5/16 were the parent, 2/16 were the sibling, and 1/16 was the cousin of the index patient. The mean age of the family representative interviewed was 43.4 years (± 9.3 SD), and they had been followed at Stanford for a mean of 7.3 years (± 4.2 SD). The majority of interviewees were White (9/16) and female (12/16), and the most common clinical indication was inherited cardiomyopathy (10/16). Full demographic information is presented in Table 1, and additional details on the structure of each family are provided in Supplementary Table 1.

**Table 1.**
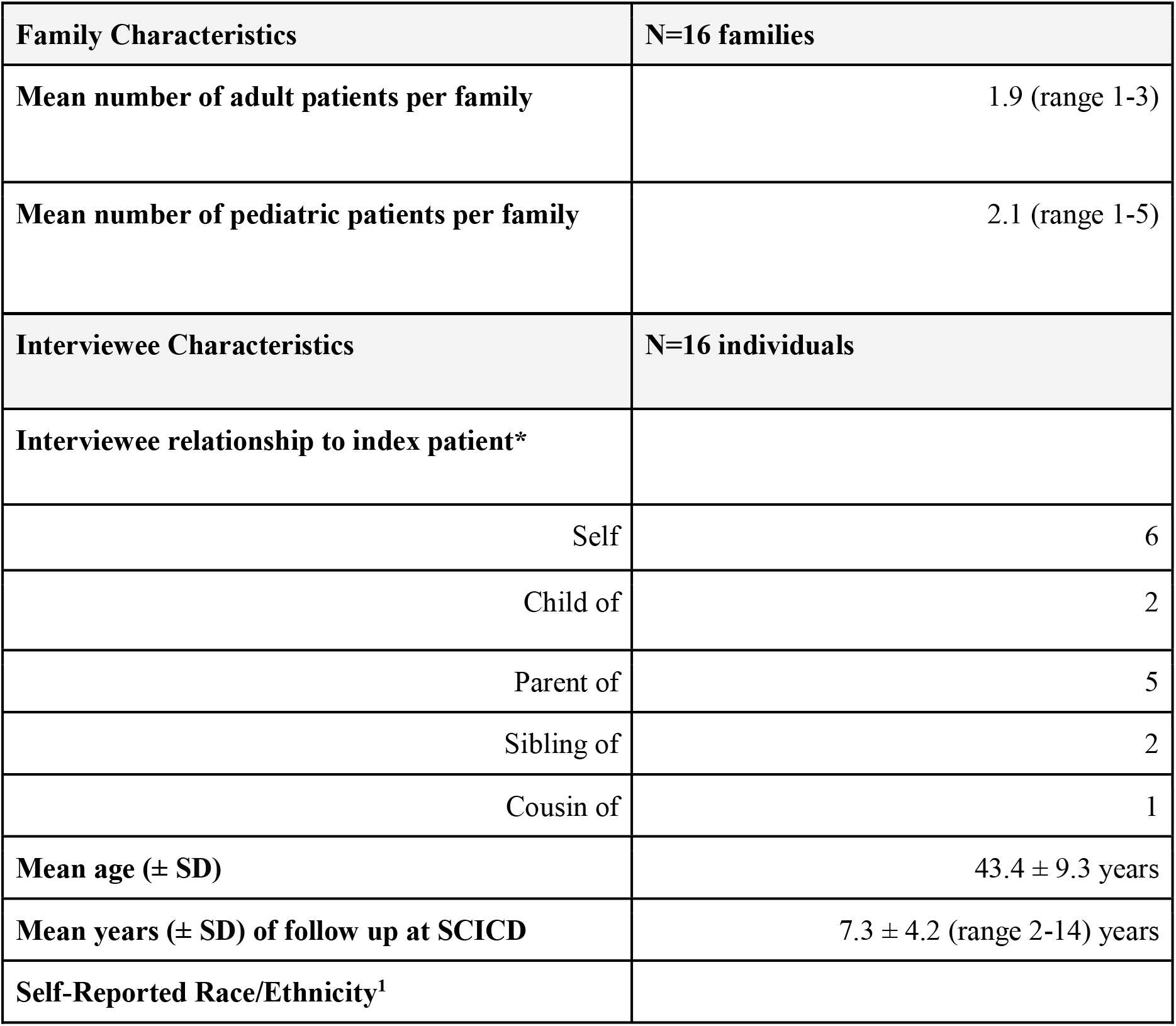
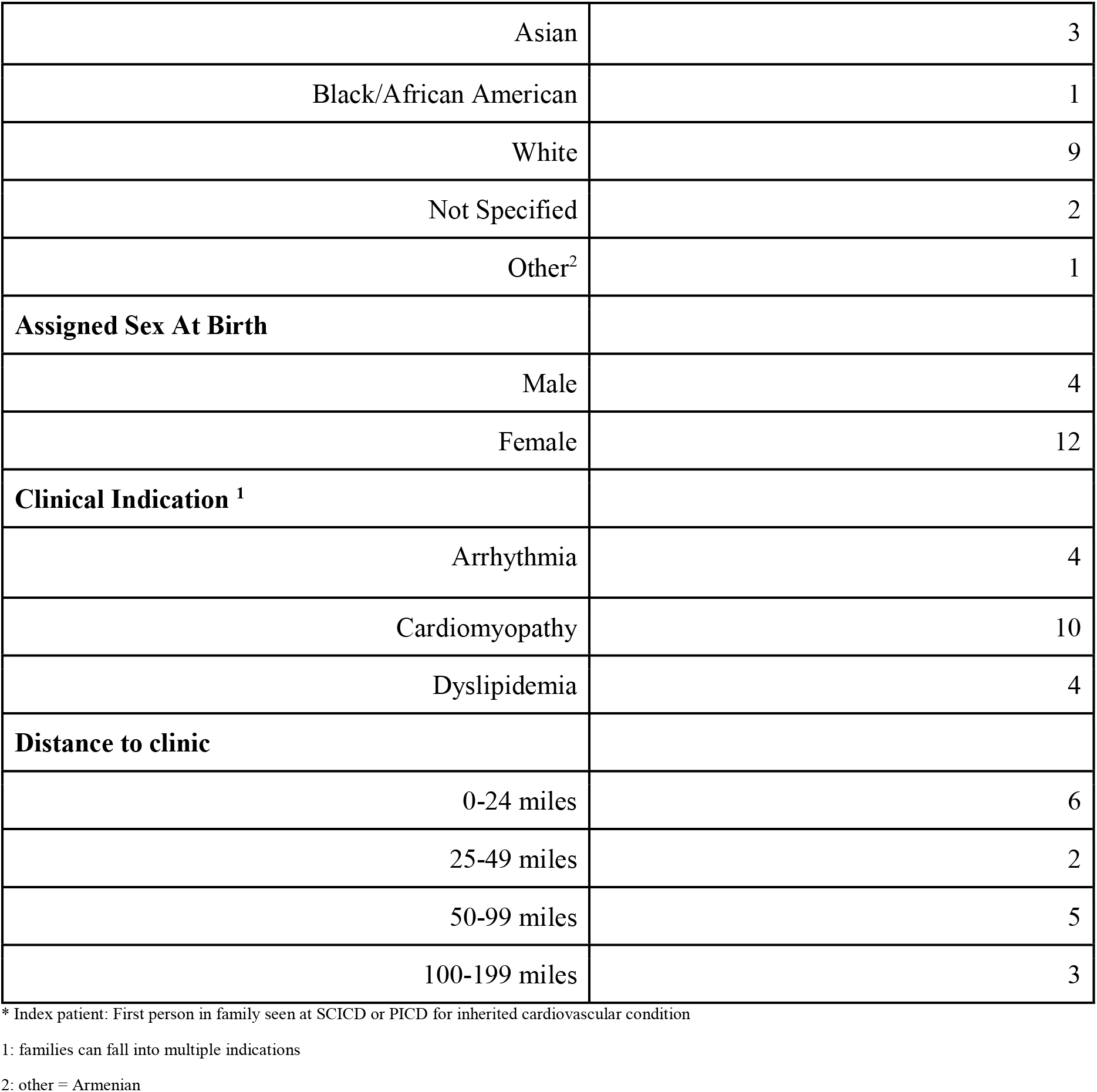
Demographics.

Participants were asked about their experience with the current clinic structure of separate pediatric and adult clinics. Interviewees ranked the ease of completing all testing (e.g. echocardiograms, EKGs, and labs) prior to their appointments as easy or very easy (Figure 1a). 75% (12/16) of families indicated they would prefer testing on the same day as their clinical appointments (Figure 1c). Of note, families who preferred to complete testing before their appointment day were enriched for dyslipidemias (75%, 3/4). The majority of families reported being either somewhat or very satisfied with information sharing between the adult and pediatric care teams (92.8%, 13/14 assessed) (Figure 1b).

**Figure 1.**
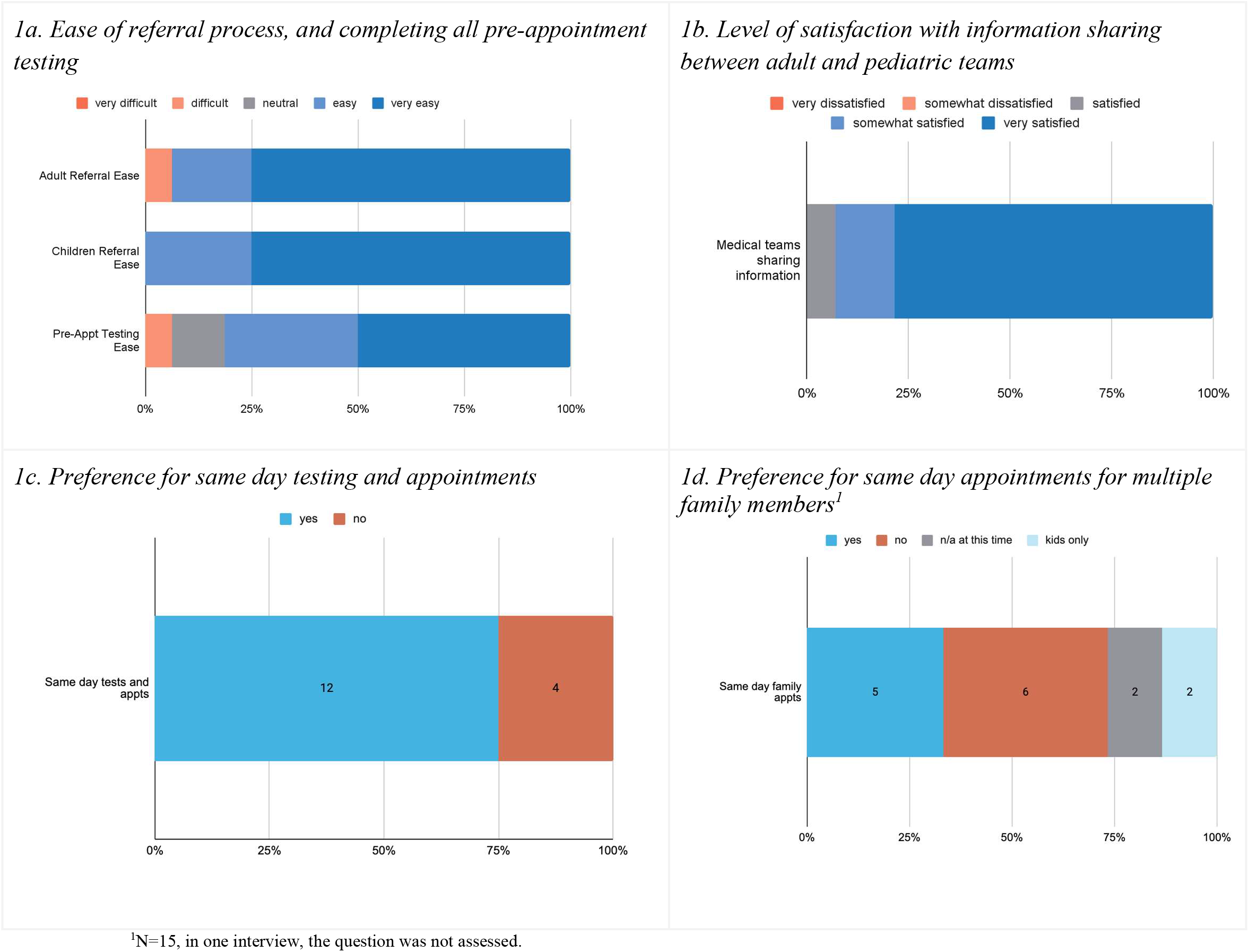
Experience with Current Clinics (n=16)

Within the current clinic structure, family preference for having same day appointments for multiple family members was varied, with 40.0% (6/15) of families not interested, 33.3% (5/15) of families interested, and 13.3% (2/15) preferring to only schedule multiple children’s appointments on the same day (Figure 1d).

Of the 16 families included, 81.2% (13/16) said they would find the option of family SMAs beneficial (Table 3). For families interested in family SMAs who opted into recorded interviews (n=12), responses were further analyzed on preferences regarding the details of such appointments. Interviewees were split on their preference for what information they would like discussed in appointments, with 50% feeling that all information would be helpful, and 50% preferring that serious information, such as information about major changes in medical status or management, would be better kept private in individual visits (Table 3). Interviewees also varied in their preferences regarding whether visits should be with immediate family members only or with extended family members, as well as their preferences for visit modality (in-person vs. virtual) (Table 3).

Two key categories were developed through content analysis: *Care Quality* (Table 2a) and *Logistics and Access* (Table 2b).

**Table 2A.**
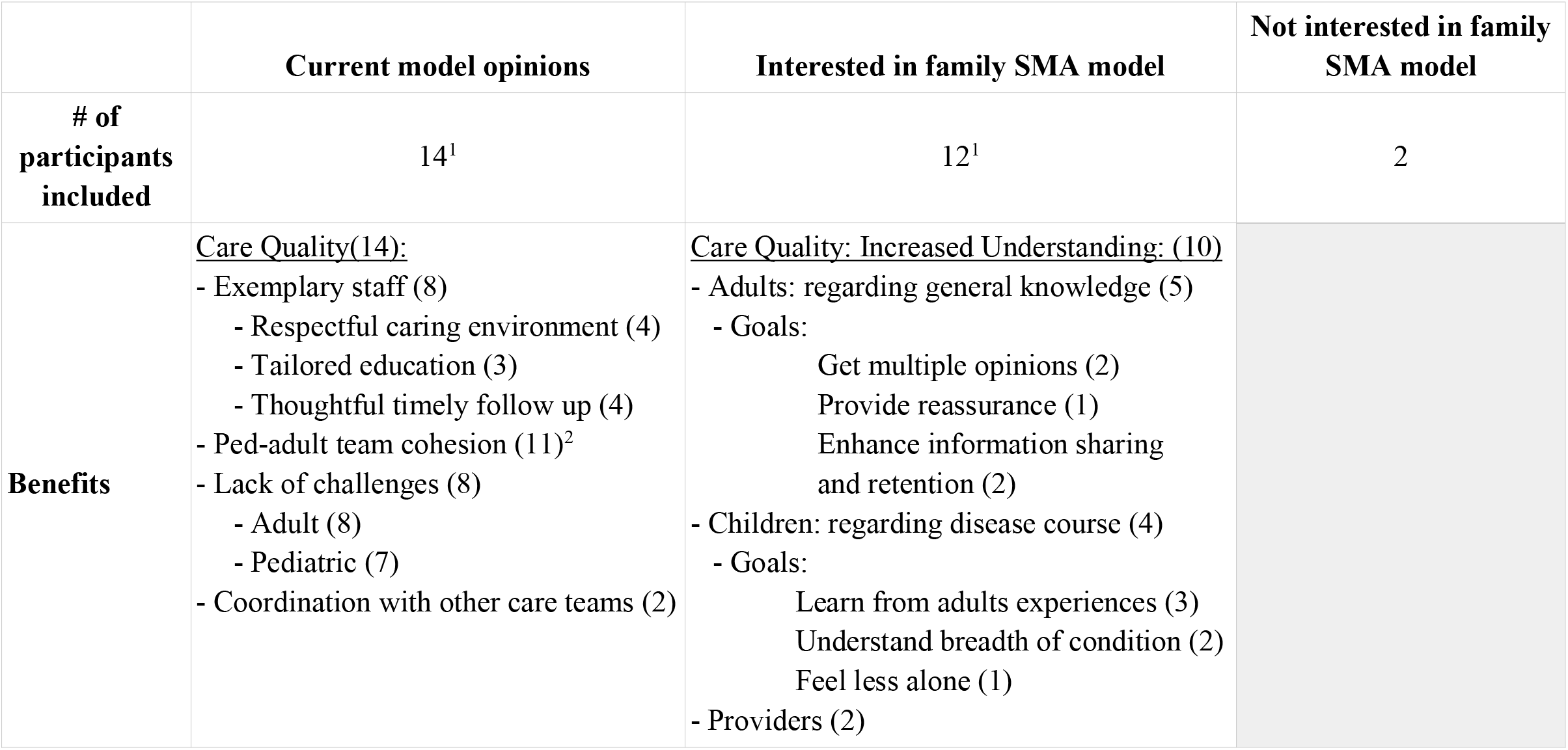
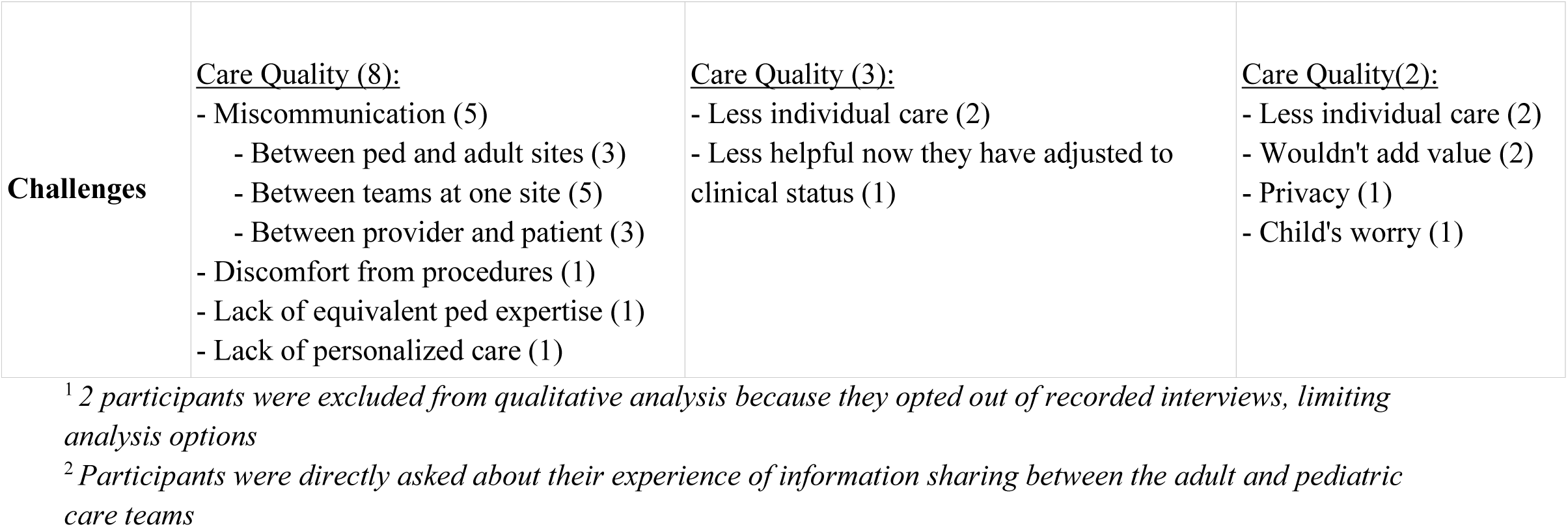
Identified Care Quality benefits and challenges of models of care.

**Table 2B.** Identified Logistical and Access-based benefits and challenges with models of care.

|  | **Current model opinions** | **Interested in family SMA model** | **Not Interested in family SMA model** |
| --- | --- | --- | --- |
| **# of participants included** | 14<sup>1</sup> | 12<sup>1</sup> | 2 |
| **Benefits** | <u>Logistics/access (12):</u><br>- Flexibility with scheduling options (4)<br>- Scheduling team communication (9)<br>- Streamlined appointment days (6) | <u>Logistics/Access (6)</u><br>- Would help with distance (4)<br>- Would help with timing (3) |  |
| **Challenges** | <u>Logistics/Access (12):</u><br>- Scheduling (10)<br>- Availability/wait times (6)<br>- Outside hospital referral (3)<br>- Challenges of clinic coordination (3)<br>- Forced rescheduling (1)<br>- Location from home (8)<br>- Distance between Stanford buildings (4)<br>- Insurance (5)<br>- Time with physician (4)<br>- EMR system (3)<br>- Billing (2) | <u>Logistics/Access (4)</u><br>- Challenge for clinic to coordinate (2)<br>- Challenge for family with timing/location (3) | None identified |

<sup>1</sup> 2 participants were excluded from qualitative analysis because they opted out of recorded interviews, limiting analysis options

### 1. Experience with current model of care

#### 1a: Current Model: Care Quality

##### I. Identified Care Quality Benefits

In regards to care quality in the current model of care, 100% (14/14) of interviewees identified at least one benefit. These benefits included exemplary staff (57.1%, 8/14) who created a respectful and caring environment (50%, 4/8), tailored education to the family (37.5%, 3/8), and thoughtful and timely follow-up (50%, 4/8) (Table 2a). For example, one participant shared:

> *“Each team member there just treats children with such care and respect… in something where there’s so much hard, and challenges going on… I’m just impressed at how they can make it such a positive experience.”*

The general lack of challenges or problems with the current model of care was also identified as a benefit in 57.1% (8/14) of interviews (Table 2a). The majority of interviewees found the ability of the pediatric and adult teams to communicate with each other to be a benefit in the current clinic model (78.6%, 11/14) (Table 2a). In describing the collaboration between adult and pediatric teams, one participant stated:

> *“The initial appointment with my children… given their ages, some were a little worried. Some don’t understand it. I’d already been going and seeing [the cardiologist], who is very informed about that gene, obviously. So when I went to the child’s physician in the child hospital, they knew about it… and kind of explained it to them as well, but in a not scary way. And with their knowledge and already knowing what was going on, it made the process for the children easy”*

##### II. Identified Care Quality Challenges

Despite the above benefits, 57% (8/14) of participants identified challenges in the current model of care (Table 2a). Miscommunication was the most common identified problem (35.7%, 5/14), between the pediatric and adult sites (60%, 3/5), different medical providers at one site (100%, 5/5), and between a provider and patient (60%, 3/5) (Table 2a).

#### 1b: Current Model: Logistics and Access

##### I. Identified Logistical and Access-based Benefits

Within the current model of care, consideration of logistics and access was a major category that emerged. Within the current model, 85.7% (12/14) of interviewees identified at least one benefit (Table 2b). The most common benefit identified was the scheduling team’s communication (64.2%, 9/14) (Table 2b). Additional logistics around scheduling were seen as a benefit, including the flexibility of scheduling options (28.6%, 4/14) and the ability to plan stacked pre-appointment testing or multiple family visits on the same day, streamlining their own appointment schedules (42.8%, 6/14) (Table 2b). Regarding streamlining appointment days, one participant shared:

> *“They would help me get the echo of the labs and the consultation all in one day. They did a really good job at helping me get it all in one day, and that way I didn’t have to drive the distance.”*

##### II. Identified Logistical and Access-based Challenges

When identifying the challenges of the current model of care, 85.7% (12/14) of interviewees identified a challenge related to logistics and access (Table 2b). Scheduling again emerged as an area of consideration for a majority of participants (71.4%, 10/14) (Table 2b). In terms of scheduling challenges, 42.9% (6/14) of interviewees had difficulties with appointment availability or wait times, 21.5% (3/14) noted difficulties with the referral process from outside hospitals, and 21.5% (3/14) had challenges with coordinating different appointments (either needing to travel to the hospital multiple times or issues with how appointments overlapped) (Table 2b).

Separately from scheduling, 57.1% (8/14) of interviewees found the distance from their homes to the clinic a challenge, and 28.6% (4/14) found the distance between the buildings they needed to access at Stanford a challenge (Table 2b).

### 2. Family SMA Model of Care

#### 2a. Family SMA Care Quality

##### I. Potential Care Quality Benefits

When asked about potential clinical benefits to the proposed family SMA, the majority of interviewees interested in such visits thought this model would help increase understanding for children, adults, and/or providers (83.3%, 10/12, Table 2a). For adult family members, increasing general knowledge about the condition was reported by 41.7% (5/12) of interviewees (Table 2a).

Increasing adult knowledge was felt to be bolstered by family SMAs in three ways. First, interviewees felt that having multiple provider opinions in the room would help answer questions from a variety of sources (16.7%, 2/12, Table 2a): *“It’d be great to get multiple opinions from different areas of expertise, just talking about everything,*” shared one participant. Secondly, participants shared that having multiple family members in the room would help with information retention for the family unit as a group (16.7%, 2/12):

> *“Honestly, it would just be really efficient for you guys and for us to tell us all the information just once and for us to all hear it just once, and then we can all ask questions and learn from each other.”*

Lastly, one interviewee shared that learning from each other’s disease course would help reassure others that there are treatment options and make others more comfortable with their own diagnosis (8.3%, 1/12):

> *“I just think it’s kind of a good idea because then you know everybody knows more about, you know, what condition you have. It’s not like a big secret, HCM. So maybe to make it more understandable that it is something that can be treated, not to worry that somebody’s going to, you know, die on you in the next second. To reassure people that there are treatment options. Not cures, but you know it’s treatable.”*

Family representatives felt that family SMAs would also increase pediatric patient knowledge of the general disease course (33.3%, 4/12, Table 2a). Family representatives felt that these visits could allow children to learn from adults’ experiences (25%, 3/12). The information families wanted shared (Table 3) was related to the goals families had for children’s learning.

**Table 3:** Preferences about family SMA details.

| **Interest In Participating in Family SMA** |  | **N=16<sup>1</sup>** |
| --- | --- | --- |
| **No** |  | **2** |
| **Yes, but children currently too young** |  | **1** |
| **Yes:** |  | **13** |
|  | **Information to Discuss in Family SMA** | **N=10<sup>2</sup>** |
|  | Everything | 5 |
|  | Privacy with serious information<sup>3</sup> | 5 |
|  | **Family Members to be Included** | **N=12** |
|  | All family members | 4 |
|  | Immediate family members only | 5 |
|  | N/A<sup>4</sup> | 3 |
|  | **Mode of Family visit** | **N=12** |
|  | In person | 6 |
|  | Telehealth | 2 |
|  | Both | 4 |

<sup>1</sup>2 participants (1 = yes, 1 = yes, but children currently too young) were only included in simple yes/no responses because they opted out of recorded interviews, limiting analysis options.
<sup>2</sup>For information to discuss in visit, n=10; 2 participants not assessed.
<sup>3</sup>Serious information was defined as sensitive information about major changes in medical status or management, immediate family was considered members of the family representative's nuclear family (spouse and children).
<sup>4</sup>Participants in the N/A category only had immediate family members who would be eligible for a coordinated family visit.

For some families, they wanted this knowledge to be reassuring:

> *“I would like both of them to talk about [how] this is a disease, not ending your life… Your mother, she got diagnosed, and your brother and your uncle, and everything will be fine. And we are now discovering new medication. And even if you are in the case that you need it, you will be in good hands. And they give him positive you know vibes for my son… And as I said, if something changes or something’s not well, I would like both of them to talk with me first.”*

A different family wanted this learning to encourage the child to take their health more seriously: “*Right now, it’s like he’s not taking it seriously, and he’s getting older now. And so I don’t know why it’s still not clicking for him. And he sees what I go through, have to go to the hospital and stay and have to get this heart transplant and all that. So yeah, I’d rather for him to hear everything [so] that he can take it seriously.*”

Additionally, families thought these visits could help children understand the breadth of how the condition presents more holistically (16.7%, 2/12), and feel less alone in their own diagnoses (8.3%, 1/12):

> *“She’d probably understand more that, you know, she’s not going through this by herself*. *You know, when she’s five, it’s going to be, you know, a lot to take in all at once that she has to go to the hospital and stuff like that. So that alone, her seeing that, you know, I also, you know, go to the doctor and sit in the office would be good.”*

Families also felt that family SMAs could allow providers to share knowledge and understand the families clinical picture more fully (16.7%, 2/12): *“I think, the shared knowledge among three clinicians is also great if they’re all there to talk it through… It’s like having a team meeting. Different perspectives [are] better than one perspective.”*

##### II. Potential Care Quality Challenges

For those interested in family SMA visits, two participants noted concerns about less individualized care in the family setting, and one participant felt that, now that they were adjusted to a stable clinical status, there would be less benefit to this visit type (Table 2a).

For participants who were not interested in the family SMA model (12.5%, 2/16), the challenges they shared were similarly about less individualized care (100%, 2/2), and not feeling that this clinic model would add value to their care (100%, 2/2), may create added worry for their child (50%, 1/2), and may bring up privacy concerns (50%, 1/2) (Table 2a). One participant shared:

> *“Given that it’s presented itself very differently across myself and my two sisters and even my niece and my nephew, I feel like that conversation would just have to still be tailored to each one of us. And it might create more confusion because it would be hard to know which things were applicable to each of us given our unique conditions and circumstances. So I guess I just feel like medicine should be more personalized.”*

#### 2B. Family SMA: Logistics and Access

##### I. Potential Logistical and Access-based Benefits

Six of twelve participants interested in having family SMA visits felt there would be logistical/access-based benefits to this new model (Table 2b). These benefits include helping with the distance to travel for visits by combining visits (33.3%, 4/12) and helping save time for families (25%, 3/12). On timing, one participant shared the model would help *“to save a lot of time for making different appointments, and we can all be there at one time to do whatever we have to do, like any testings or checkups and all that stuff.”*

##### II. Potential Logistical and Access-based Challenges

A few participants interested in family SMA still shared concerns about logistics and access in this new model (Table 2b). This included concerns about the hospital’s ability to coordinate such visits (16.7%, 2/12), and difficulty with navigating the timing or location of these visits (25%, 3/12). One participant shared concern with timing, stating:

> *“We don’t really understand what the optimal length of appointment for a group of three people or two patients might look like versus three patients. You know If it’s just like me coming in, for example, my maintenance check every year is fast, quick, done, because your stats look okay, and you’re trending correctly. But it’s like a group setting that you go through every single person and someone that has more severe health, then maybe you have to spend more time on that. So it kind of like veers the conversation.”*

Of note, neither of the two participants who were uninterested in the family SMA model expressed concerns about the care model regarding access or logistics (Table 2b).

## Discussion

The SMA model aims to improve access to care and enhance patient education and self-efficacy. Research on SMA visits for families with inherited cardiovascular disease is limited to a single study,^17^ and to our knowledge there has been no research on incorporating the values of FCC directly into a family SMA model of care for inherited disease. For families with inherited cardiovascular conditions, coordinating care for multiple family members and ensuring the whole family has a comprehensive understanding of their condition can be challenging. Therefore, we investigated family perspectives on family SMA to help address potential unmet needs. Results show that the majority of participants felt this clinic model could add value to their care, but that there are several variables that need to be taken into consideration when thinking about future implementation as well as which families may benefit the most.

Participants in this study noted care quality benefits to the current model, but thought that family SMAs might bring added benefits such as improved education and information retention. For instance, families shared that this model would allow for input from multiple providers simultaneously and provide multiple family members the opportunity to listen to this information, allowing for cohesive messaging and leading to an increase in overall retention. For children, adult family representatives shared that an added educational benefit may be the opportunity to learn from their adult family members’ clinical experiences and better understand the breadth of the condition. Increased understanding has been previously reported as a benefit of family centered care, with improvements in both knowledge and comprehension of a variety of indications ranging from diabetes self-management to nutrition.^5^ Similar benefits have been shown in research on genetic counselors’ experiences conducting family group appointments for hereditary cascade testing, where genetic counselors reported that shared appointments helped prevent information loss in translation between family members and fostered mutual understanding of information shared by providers.^14^

Participants in the present study also felt that this model may help adults normalize the condition amongst themselves and provide reassurance to family members. Additionally, they felt this would support children by making them feel less alone in their own diagnoses. Research on other SMA use shows patients reported benefits for normalization and reassurance from their peers in a systematic review study.^12^ Support groups for genetic conditions have also been shown to have benefits similar to the family SMA care quality benefits identified, such as helping patients better understand their conditions and connecting them with others who have similar lived experiences.^21^ These findings show family SMAs may hold the most value for families with a less cohesive understanding of their condition, and who are looking to form a sense of togetherness in navigating their care.

Despite these benefits, participants also expressed some concerns that the family SMA model allowed for less individualized care, potential privacy concerns, and some saw limited added value altogether. Individualized care concerns have not been well represented in previous research; instead, studies of SMA have shown that patients enjoy learning from other patients’ experiences and questions.^22^ Interestingly, the study by Chang et al, which assessed cascade genetic testing via family group appointments, found that genetic counselors believe the model of care is self-selecting, with closer-knit families more interested in the group setting.^14^ This suggests that family SMA may potentially better support certain family dynamics over others.

From an access and logistics perspective, families felt that the family SMA model could provide additional benefits in reducing travel, but may pose new coordination challenges for themselves and the clinic. Families felt that these joint visits could lead to fewer trips, address the challenge of distance to the clinic, improve visit timing, and overall provide the benefit of getting everything done at once. Logistically, some families were concerned that the visit day may pose a challenge for both themselves and the clinic to coordinate. Some families were concerned about whether this appointment modality may be considerably longer, and some were concerned the clinic may not be able to effectively streamline a joint appointment day. These are reasonable considerations, as research on administrator perspectives of SMAs identified logistical barriers such as physical space, administrative time, and hospital buy-in.^12^

For families interested in participating in a family SMA, there was variation in communication preferences. Families were split in their opinions about what information to discuss, with half preferring that communication be fully open, and half preferring that serious information be discussed privately in an individual setting. Confidentiality and privacy concerns have been reported as barriers to SMA in previous research.^12^ However, some studies in the primary care and pulmonary medicine setting have shown that these concerns dissipate after experiencing a SMA firsthand.^22,23^ This research contrasts, however, with genetic counselor experiences with family group appointments for cascade testing, where many had encountered a patient who was not comfortable with completely open discussion.^13^ This indicates that introducing the familial element to an SMA model has unique considerations for openness with information sharing, and suggests that direct discussion with affected family members about their preferences around sensitive information and privacy is needed to set expectations and help guide the content of the visit.

Research on communication about inherited genetic conditions with children shows that parents can find discussions about future risk very difficult and emotionally painful, and that they often delay these disclosures until a major medical event has occurred ^24^ One avenue of further investigation might be whether a family SMA might provide a safe opportunity for families to include their children in difficult discussions and provide a developmentally appropriate opportunity for their learning. One study evaluating potential benefits of multi-family discussion groups for inherited genetic conditions found that the main benefit would be empowering families to have conversations with their children. More research is needed to better understand the factors that influence families’ communication preferences.

It is also important to recognize that inherited cardiovascular diseases are a heterogeneous group, and there may be indication-specific considerations important for family SMA implementation. For example, in this study, most participants preferred to complete pre-visit testing on the same day as their appointment; however, of the four families who preferred separate pre-visit testing, three were seen for dyslipidemia. The pre-visit workup for dyslipidemias involves lab testing to evaluate lipid levels, which takes time to yield results and is important for clinical decision-making.^25^ Recognizing that inherited cardiovascular diseases have differences in treatment and management is important, as these differences should be considered when assessing family needs and clinical logistics. These differences may also impact the clinical utility of a family SMA model. For patients with arrhythmias, assessing multiple family members all at one time with EKGS may help clarify ambiguous genotype/phenotype data in real time and provide important information for disease diagnosis and management,^26^ making these conditions particularly suited for family SMA. Based on this data, family SMA implementation may benefit from including clear pre-visit contracting about who to include, privacy and sensitive discussion expectations, and condition-specific visit logistics.

This study was limited to perspectives on perceived benefits and challenges of family SMAs rather than a direct assessment of patients who had undergone such visits, as this research was conducted prior to the clinic rollout. The small sample size was a limitation, and non-White and male family representatives were underrepresented. Additionally, we are aware that there can be insurance barriers that allow only pediatric patients to be seen at the institution, and so families where adult patients were unable to access care were not represented. The families in the study were varied in family structure, but do not represent all family structures and complex family dynamics. Family representatives were limited to adult patients, and so the direct perspectives of pediatric patients were not included. Further research could explore children’s perspectives on the value of family care visits directly. Interviews were limited to families, and so the opinions of key stakeholders like clinical providers and hospital administration is not represented by this data. Additional research with these stakeholders could help clarify specific staffing needs for family SMA visits and integration with existing hospital workflows.

## Conclusion

In sum, this study found that the majority of families believed a family SMA model would provide added benefit to families with inherited cardiovascular disease, but family-specific needs must be thoughtfully taken into consideration. The family SMA model may benefit from being used as an additional option for families who have specific care needs, rather than a replacement of the current clinic model. Future research focused on assessing patient outcomes following a family SMA is recommended to evaluate how the implementation of family SMAs impacts family-centered care experiences.

## Data Availability

Data is available upon request.

## Acknowledgements

The research team gratefully acknowledges the patients and families for their time and openness in sharing their perspectives, and the Stanford Center for Inherited Cardiovascular Disease, in particular Kyla Dunn, Joshua Knowles, Chloe Reuter, and Ryan Murtha, in conceptualizing this project and providing input on the interview guide. This work was supported by Stanford University’s Genetic Counseling Program.

